# SARS-CoV-2 sero-surveillance after the first and second waves of the epidemic in Panama

**DOI:** 10.1101/2025.07.20.25331847

**Authors:** Carlos Lezcano-Coba, Josefrancisco Galue, Charles Whittaker, Cathal Mills, Rodrigo de Antonio, Xavier Saenz-llorens, Luis Felipe Rivera, Xacdiel Rodriguez, Danilo Franco, Arturo Rebollón, Edward Espinosa, Catherine Castillo-Castillo, Alexander Martínez, Mabel Martínez, Xenia León, Adelain Ríos, Joseph Arauz, Publio Gonzalez, Claudia Dominguez, Sandra López-Vèrges, Bernardo González, José Jimeno, Anayansi Valderrama, Ingrid Murgas, Kathryn A. Hanley, Nikos Vasilakis, Ilaria Dorigatti, Michaela Bueneman, Nuno R. Faria, Moritz Kraemer, Blas Armien, Christl A. Donnelly, Jean-Paul Carrera

## Abstract

**Introduction:** In the context of the ongoing global SARS-CoV-2 pandemic and potential future viral epidemics, seroepidemiological studies are essential for understanding epidemic dynamics. Panama, a country with a strategic geographic position that serves as a regional hub for international travel and commerce. As such, understanding local transmission patterns is critical for developing evidence based on interventions that can effectively anticipate and mitigate emerging viral threats.

**Methodology:** We conducted a population-based cross-sectional study in the provinces of Panamá and Panamá Oeste, targeting the 10 townships with the highest cumulative COVID-19 incidence. A multi-stage cluster sampling strategy was employed using the 2010 National Housing Framework. The first survey was conducted from November 30 to December 4, 2020. A follow-up survey was carried out from June 14 to July 10, 2021, to assess seroconversion and seroreversion. Serum samples were analyzed using two electrochemiluminescence immunoassays (Cobas and Vitros) to detect SARS-CoV-2-specific IgG, IgM, and IgA. We estimated the effective reproduction number (Rt) and fitted modified Poisson regression models to identify risk factors associated with seropositivity.

**Results:** A total of 2198 participants were recruited in the first round, and 547 were successfully followed up in the second round. SARS-CoV-2 seroprevalence increased from 24.7% (95% CI: 23.0–27.0%) in the first round to 66.2% (95% CI: 62.0 - 70.0%) in the second. The seroconversion rate was 42.9% (95% CI: 38.0 - 47.0%), while seroreversion was rare (0.9%; 95% CI: 0.3 - 2.0%). The most parsimonious multivariable model identified Indigenous ethnicity, contact with a confirmed case, cohabiting with an infected household member, and prior COVID-19 diagnosis as significant risk factors for seropositivity. In contrast, higher education, belonging to other ethnic groups, and consistent mask use at work were protective factors.

**Conclusions:** SARS-CoV-2 transmission persisted in Panama despite strict public health interventions. Household transmission, particularly among Indigenous and socioeconomically vulnerable populations, was a major driver of infection. Higher education and adherence to preventive behaviors emerged as protective factors. These findings emphasize the importance of targeted, equity focused strategies to strengthen epidemic control in Panama and comparable settings.

## Introduction

SARS-CoV-2, the etiologic agent of COVID-19, was first identified in late 2019 in Wuhan, Hubei province, China, and rapidly spread globally during early 2020^1,2^. By September 2024, approximately 776 million confirmed cases and over 7.1 million deaths had been reported worldwide^3,4^. In this context, sero-surveillance plays a key role in understanding the true extent of viral spread, the dynamics of transmission, and identifying populations at increased risk of infection. Such insights are essential for guiding public health interventions, evaluating the impact of control measures, and improving preparedness for future outbreaks.

Panama reported its first confirmed imported case of SARS-CoV-2 on March 9, 2020, from a traveler arriving from Spain^5^. The following day, a fatal infection in an individual with no travel history was confirmed as SARS-CoV-2. Genomic epidemiology surveillance suggests that the A.2.1/19B variant, originating from Spain, had already been circulating in Panama since early February 2020^5,6^. On June 1, 2020, Panama implemented control measures outlined by the Ministry of Health (MINSA) in Resolution No. 1420, based on guidance from the World Health Organization (WHO) and the Centers for Disease Control and Prevention (CDC) ^7,8^. These measures included lockdown, the use of masks, social distancing, hand hygiene, and the enforcement of cluster quarantines^7^.

Restrictions in Panama remained in place for six months ^9^. Despite the implementation of rapid and extended control strategies, Panama exhibited a high early transmission potential, with an estimated basic reproduction number R_0_ of 2.22 (95% CI 2.08–2.37), an exponential growth rate of 0.13 per day, and a doubling time of 7.7 days, leading to a widespread epidemic across the country^5^. By early July 2020, daily confirmed cases had reached 1,000, and by mid-December 2020, they exceeded 3,000 per day^10^. As of September 2024, Panama has reported a total of 1.04 millions confirmed SARS-CoV-2 infections and over 8754 deaths due to COVID-19^10^.

Previous seroprevalence studies in Panama have been conducted among blood donors and healthcare workers, estimating an overall seroprevalence of 11.6% (95% CI: 8.5–15.8%)^11^. However, population-based seroprevalence estimates are essential to achieve a more comprehensive understanding of transmission dynamics, to identify vulnerable populations, and to inform more effective control strategies. Panama continues to report SARS-CoV-2 cases, with sporadic outbreaks occurring in various regions of the country, underscoring the ongoing importance of epidemiological and serological surveillance. In this context, we present the findings of a cross-sectional, multi-stage cluster-based serosurvey conducted in 2020, along with a risk factor analysis, carried out after the first and second epidemic waves in ten townships with the highest incidence rates of clinically confirmed COVID-19 cases in the provinces of Panamá and Panamá Oeste.

## Methods

### Ethics statement

The seroprevalence study was conducted as part of the national pandemic response to SARS-CoV-2 in the provinces of Panamá and Panamá Oeste. It adhered to ethical guidelines for human research established by the 18th World Medical Assembly in Helsinki in 1964 and its subsequent revisions. The study also complied with Panama’s national and local ethical requirements, including those related to ethical review, informed consent, and regulations designed to protect participants’ rights and welfare in epidemiological and biomedical research. The study protocol was reviewed and approved by the Panamanian Ministry of Health [1522] and the National Bioethics Committee [PT-023]. All participants provided written informed consent, and confidentiality was ensured by removing any identifying information.

### Epidemiological investigation

COVID-19 cases and deaths confirmed data for Panama, covering the period from March 2020 to December 2021, were obtained from the Ministry of Health. We estimated key epidemiological parameters, including the effective reproduction number (R_t_), to provide a comprehensive overview of the country’s epidemiological status. Additionally, we summarized both non-pharmaceutical and pharmaceutical interventions implemented in Panama during the study period to contextualize the measures adopted in response to the outbreak.

### Study site

The provinces of Panamá and Panamá Oeste are situated in the southern part of Panama. Panamá province has a total population of 450,302 inhabitants and is divided into 26 townships^13^. Panamá Oeste province has a population of 199,472 inhabitants and is composed of 60 townships ^13^. Both provinces feature a tropical climate with two well-defined seasons: the rainy season and the dry season. Temperatures typically range from 20 to 34 °C, while the average annual precipitation is around 1900 mm (Fig. 1A)^14^.

**Figure 1.**
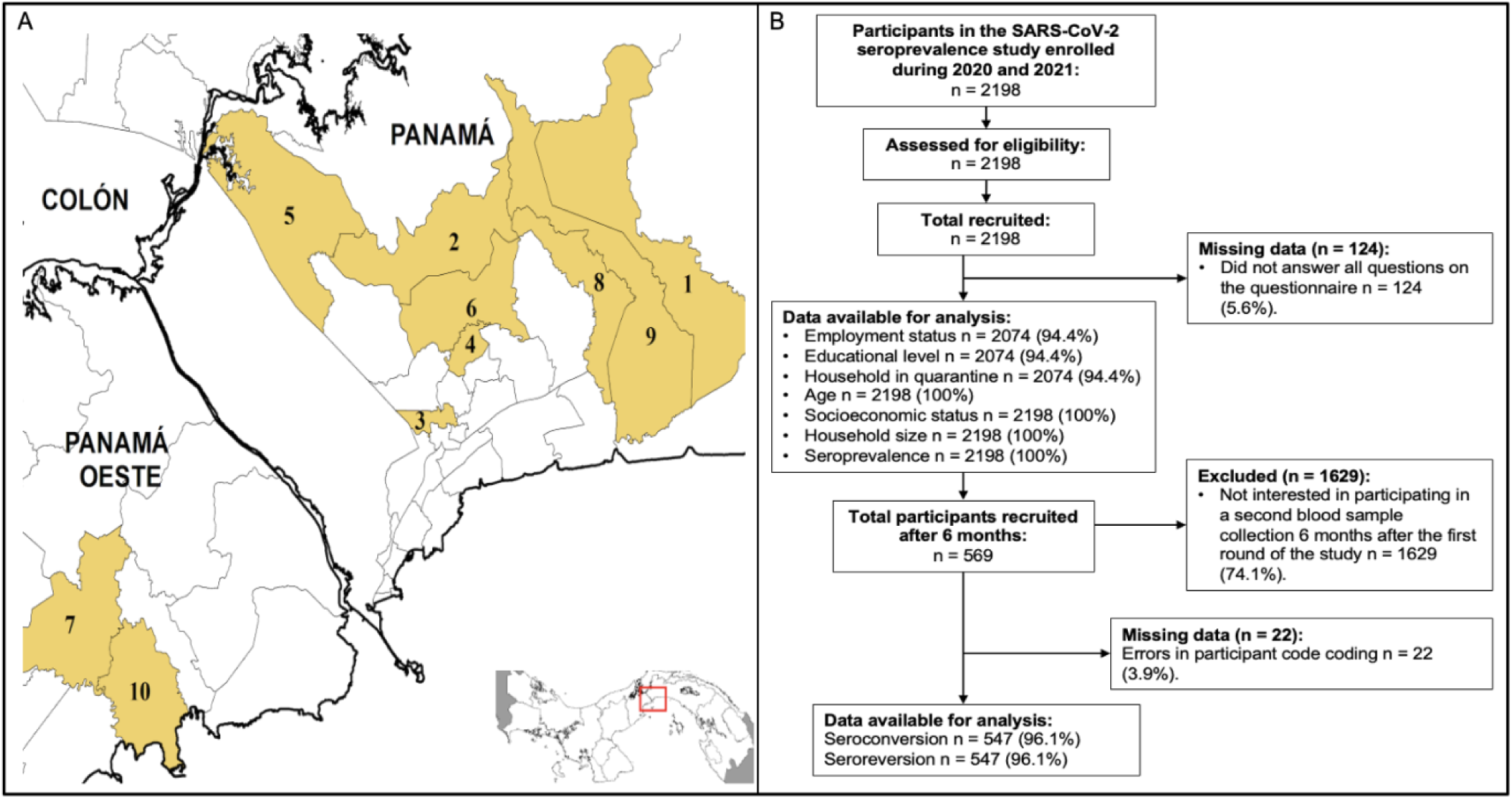
A) Map of Panama and study site. **The following districts were selected as the study site:** 1-24 de Diciembre, 2-Alcalde Díaz, 3-Amelia Denis de Icaza, 4-Belisario Frias, 5-Chilibre, 6-Ernesto Córdoba Campos, 7-Juan Demóstenes Arosemena, 8-Las Mañanitas, 9-Tocumen y 10-Vista Alegre**. B) Participant enrollment flowchart during the Seroprevalence study in Panama.** Flowchart of study participants in the seroprevalence study conducted in the provinces of Panama and West Panama during the years 2020 and 2021.

### Study design

A population based cross sectional study was undertaken after the first epidemic wave in Panama from November 30 to December 4, 2020, to assess the prevalence of antibodies to SARS-CoV-2. The sampling frame used for this study was the 2010 National Housing Framework of the National Institute of Statistics and Census (INEC) of Panama, constructed from cartographic information obtained from the last National Population and Housing Census in 2010 and demographic estimates by age in five-year periods as of March 31, 2020, in the provinces of Panama and Panama West. Multistage cluster sampling was undertaken at each township. A follow up of all original participants recruited during November 30 to December 4, 2020, was attempted after the second epidemic wave from June 14 to July 10, 2021 (Fig. 1B, 2B).

### Recruitment

The epidemiological survey,using probabilistic sampling, was structured to yield independent results for each of the following study domains: Townships with higher incidence of cases and fatality rates in Panamá and Panamá Oeste Provinces.

### Sample collection

Blood sampling was carried out by appropriately trained healthcare personnel. Once participants completed the informed consent and survey, the sample collection process commenced. Participants were asked to sit on a stable surface or chair for blood collection. Trained personnel identified the vein for blood extraction, cleansed the area with a cotton swab and alcohol, and then applied a tourniquet on the participant’s arm to introduce a vacutainer needle into the vein. Subsequently, the samples were stored in coolers with cold packs at a temperature of 20°C until transportation. The amount of blood sample varied based on age: Children under 5 years old: 1 tube of 5 ml was drawn, depending on the child’s weight and general health status. For those over 5 years old: 2 tubes of 5 ml were drawn, considering the participant’s weight and overall health.

### Laboratory methods

The serum samples were analyzed using two different electrochemiluminescence tests, the commercial kit COBAS – Elecys Anti-SARS-CoV-2 N protein sensitivity of 98.8 % (CI 95 %: 98.1% - 99.3 %) and a specificity of 99.98 % (CI 95 %: 99.9% - 100.0 %) ^15^ and commercial kit VITROS - VITROS Immunodiagnostic Products Anti-SARS-CoV-2 Total Reagent Pack to detect reactive immunoglobulin G, M, A antibodies (IgG, IgM, IgA) to the SARS-CoV-2 S1 protein sensitivity less than 8 days 80.0% (56.3% - 94.3%), more than 8 days 100.0% (92.7% - 100.0%) and a specificity 100% (95% CI: 99.1% - 100.0%) ^16^. Both tests when compared to the Gold Standard (PCR). Additional information about the kits used can be found in the supplementary material.

### Statistical methods

To characterize the epidemiological dynamics of SARS-CoV-2 during the study period, national data on confirmed case incidence from March 2020 to December 2021 were analyzed. The estimation of the time-varying effective reproduction number (Rt) was conducted using a Bayesian approach implemented in the EpiEstim package in R. This model enables the estimation of Rt over time by incorporating daily incidence data, weighted according to the distribution of the serial interval of the virus, defined through an infectivity function (wₛ). The Rt estimates were based on the weighted sum of incident cases up to time t−1, allowing for the inference of changes in viral transmissibility throughout the study period (Fig. 2A).

**Figure 2.**
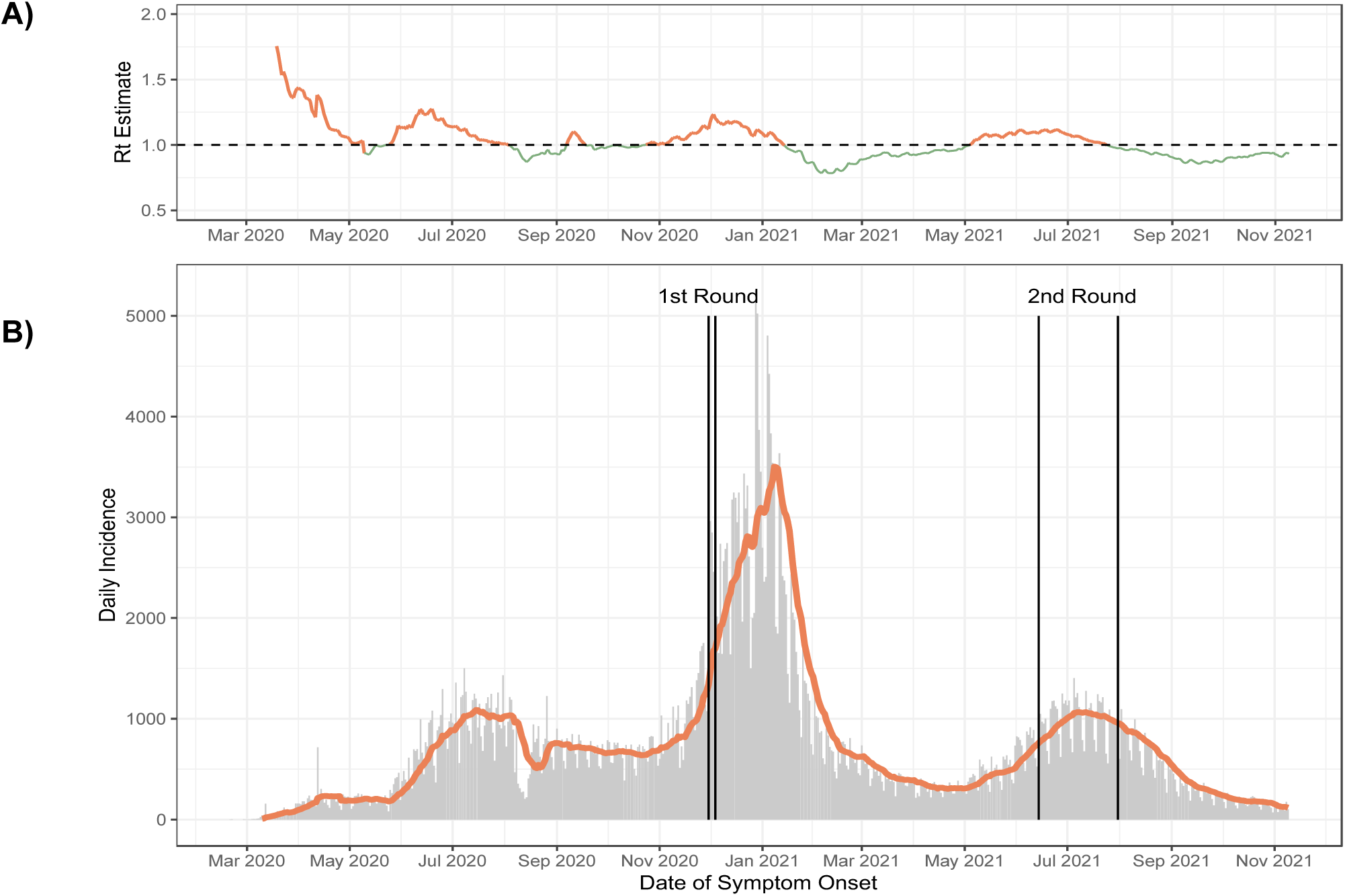
Epidemiological situation of COVID-19 in Panama, national-level case report from March 2020 to November 2021. A) Time-varying reproduction number was estimated from the epidemiological data of reported cases from March 2020 to November 2021. B) First and second rounds of SARS-CoV-2 seroprevalence study.

Descriptive statistics as absolute frecuency and percentaje were used to compare the participants characteristics and household characteristics. We report crude model and adjusted model estimates using township, weighted to match the population structure of each township, by age and sex (b) clustering at the block level. Weights (expansion factors) were provided by the Institute of National Statistical and Census (INEC) of Panama, using previously established methodology.

Frequencies and percentages were used to summarize the characteristics of the study population. The outcome variable was defined as the presence or absence of SARS-CoV-2 antibodies, based on the results of both the Cobas and Vitros electrochemiluminescence tests.

A participant was considered seropositive if one or both tests yielded a positive result, and seronegative only if both tests were negative. Seroconversion in the population was estimated using participants who were seronegative by both the Cobas and Vitros assays in the first round of the study and seropositive by both assays in the second round. Conversely, seroreversion was evaluated among participants who were seropositive by both Cobas and Vitros assays in the first round and seronegative by both assays in the second round. Independent variables include: sex, age groups, ethnicity, Educational level, Household size, Monthly household income, Township, Quarantined housing, Time living in this location, Currently working, Contact with confirmed case, Travelled outside the country, Travelled to the countryside, Family member diagnosed with COVID-19, Complies with sanitary measures, Complies with safety measures with sick family member, Wears a mask when going out, Wears a mask at work, Wears face protection, Handwashing, Gloves, Sample for COVID-19 diagnosis and Previously diagnosed with COVID-19.

To evaluate risk factors, we conducted univariate analyses using a modified Poisson regression analysis to generate more accurate estimates due to the high prevalence, with the outcome and each independent variable. For multivariable analysis, variable selection was undertaken according to relevance and significance in a stepwise fashion using the log-likelihood ratio test. The associations between each outcome and independent variables were expressed as prevalence ratios (PRs) to avoid risk inflation. Univariable and multivariate PRs and 95% CIs were calculated using the survey function to correct the study design effect and adjusting by cofounding variables sex, age groups and educational level. A threshold of 10% was established for the variance inflation factor to identify multicollinearity among the variables included in the final model. Statistical analyses were conducted using the RStudio version 4.3 and Stata version 17 MP package. A p-value <0.05 was considered statistically significant.

## Results

### Characteristics of the COVID-19 epidemic in Panama and non-pharmaceutical interventions

The first case of COVID-19 in Panama was reported on March 9. Non-pharmaceutical interventions (NPIs) were adopted on March 23, 2020, with physical distancing, hand hygiene, and the mandatory use of face masks being the first actions implemented (Fig. 3).

**Figure 3.**
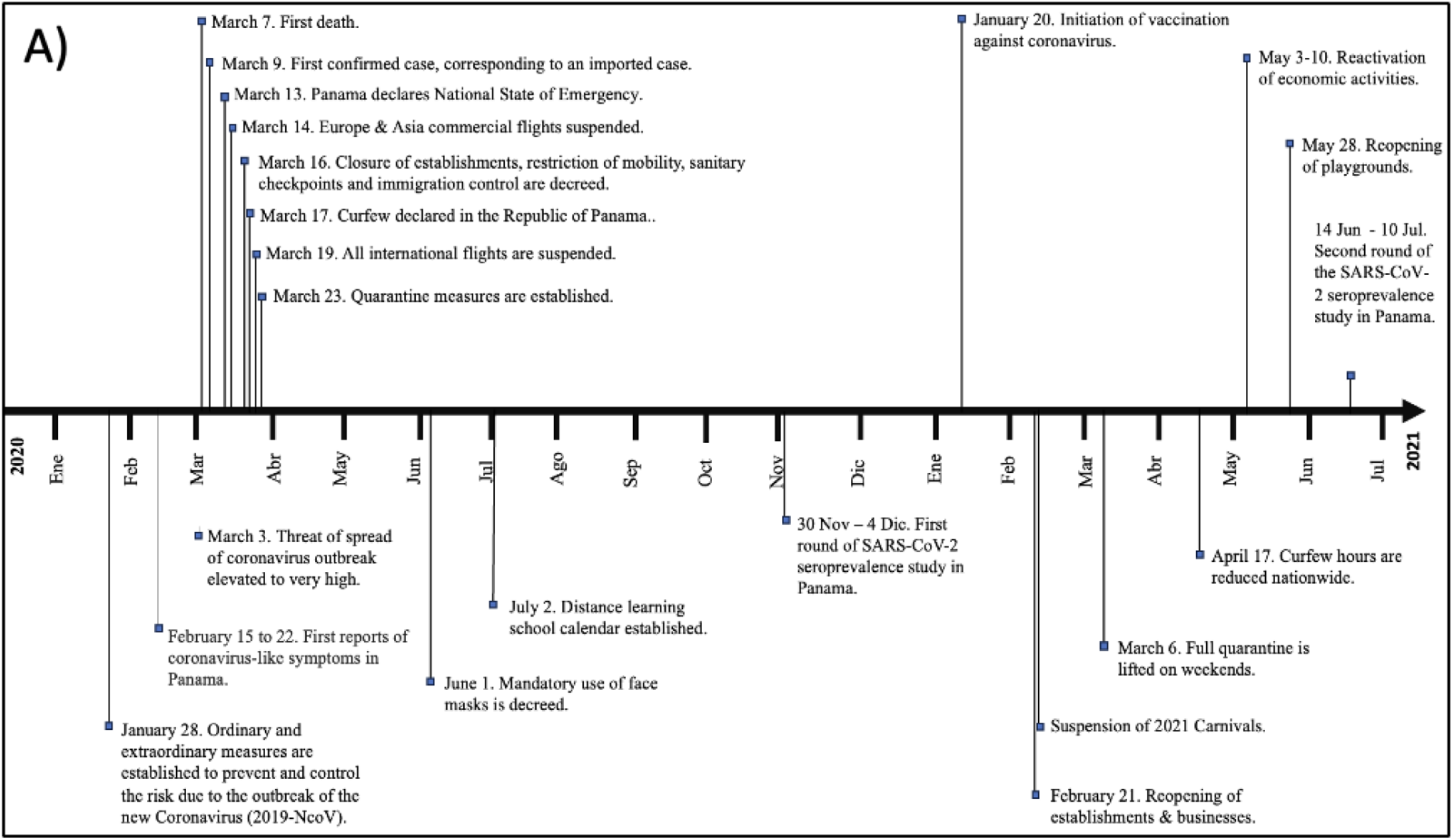
Timeline of important events recorded for SARS-CoV-2 in Panama during the years 2020 and 2021. Timeline showing the history of events related to SARS-CoV-2 transmission and control since its introduction to Panama in 2020 until the latest national seroprevalence study conducted in 2021.

The time-varying effective reproduction number (R_t_) estimates were greater than 1 during the first and second rounds of the study (Fig. 2A).

### Characteristics of the study population

From November 30 to December 4, 2020, 2198 participants were recruited in the 10 townships with the highest SARS-CoV-2 incidence rates in Panama and Panamá Oeste province^17^. We successfully followed up 547 participants in a second study round from July 14 to June 31, 2021 (Fig. 1B).

During the first round, participant ages ranged from 2 to 106 years. Were adults over 60 years represented the 24.8%. The women comprised 59.7 % of the sample, and 63.7 % of the sample were of mixed-race descent. We estimate the SARS-CoV-2 seroprevalence was 24.7% (95% CI: 23.0% - 27.0%) for the first round and 66.2% (95% CI: 62.0%–70.0%) for the second round of the study. The proportion of individuals who seroconverted seven months after the first round was 42.9% (95% CI: 38.0% - 47.0%), whereas the proportion of individuals who experienced seroreversion was 0.91% (95% CI: 0.3%–2.0%). The rest of the population characteristics are shown in (Supplementary Table 1).

### Risk factors

The prevalence of antibodies to SARS-CoV-2 studied during the first round was statistically significant, at the univariate level, for the following risk factors: belonging to an indigenous ethnicity (PR = 2.18; 95% CI: 1.53 - 3.12, p < 0.001), belonging to a low socioeconomic status (PR = 1.55; 95% CI: 1.16 - 2.08, p = 0.003), having a house in quarantine (PR = 2.36; 95% CI: 1.71 - 3.25, p < 0.001), having contact with a confirmed case (PR = 3.02; 95% CI: 2.45 - 3.74, p < 0.001), living with a family member diagnosed with COVID-19 (PR = 3.44; 95% CI: 2.76 - 4.28, p < 0.001), undergoing sample collection for COVID-19 diagnosis (PR = 2.17; 95% CI: 1.74 - 2.72, p < 0.001), and having previously been diagnosed with COVID-19 (PR = 3.81; 95% CI: 3.17 - 4.58, p < 0.001).

Meanwhile, the following were identified as protective factors against SARS-CoV-2 seropositivity: being aged over 60 years (PR = 0.71; 95% CI: 0.51–0.99, p = 0.043), belonging to other ethnic groups (PR = 0.39; 95% CI: 0.17 - 0.88, p = 0.023), and not having a family member ill due to COVID-19 (PR = 0.48; 95% CI: 0.38–0.62, p < 0.001).

The final multivariable adjusted model selected shows that being of indigenous descent (PR = 1.98; 95% CI: 1.28 - 2.93; p = 0.002), having contact with a confirmed case (PR = 1.46; 95% CI: 1.09 – 1.95; p = 0.010), living with a family member diagnosed with COVID-19 (PR = 1.98; 95% CI: 1.45 - 2.70; p < 0.001), and having previously been diagnosed with COVID-19 (PR = 2.29; 95% CI: 1.75 - 3.00; p < 0.001) were risk factors for SARS-CoV-2 seropositivity.

In contrast, belonging to other ethnic groups (PR = 0.28; 95% CI: 0.11–0.80; p = 0.016), having higher education (PR = 0.52; 95% CI: 0.36 - 0.76; p = 0.001), and wearing a mask at work (PR = 0.52; 95% CI: 0.32 - 0.85; p = 0.008) were protective factors (Fig.4).

**Figure 4.**
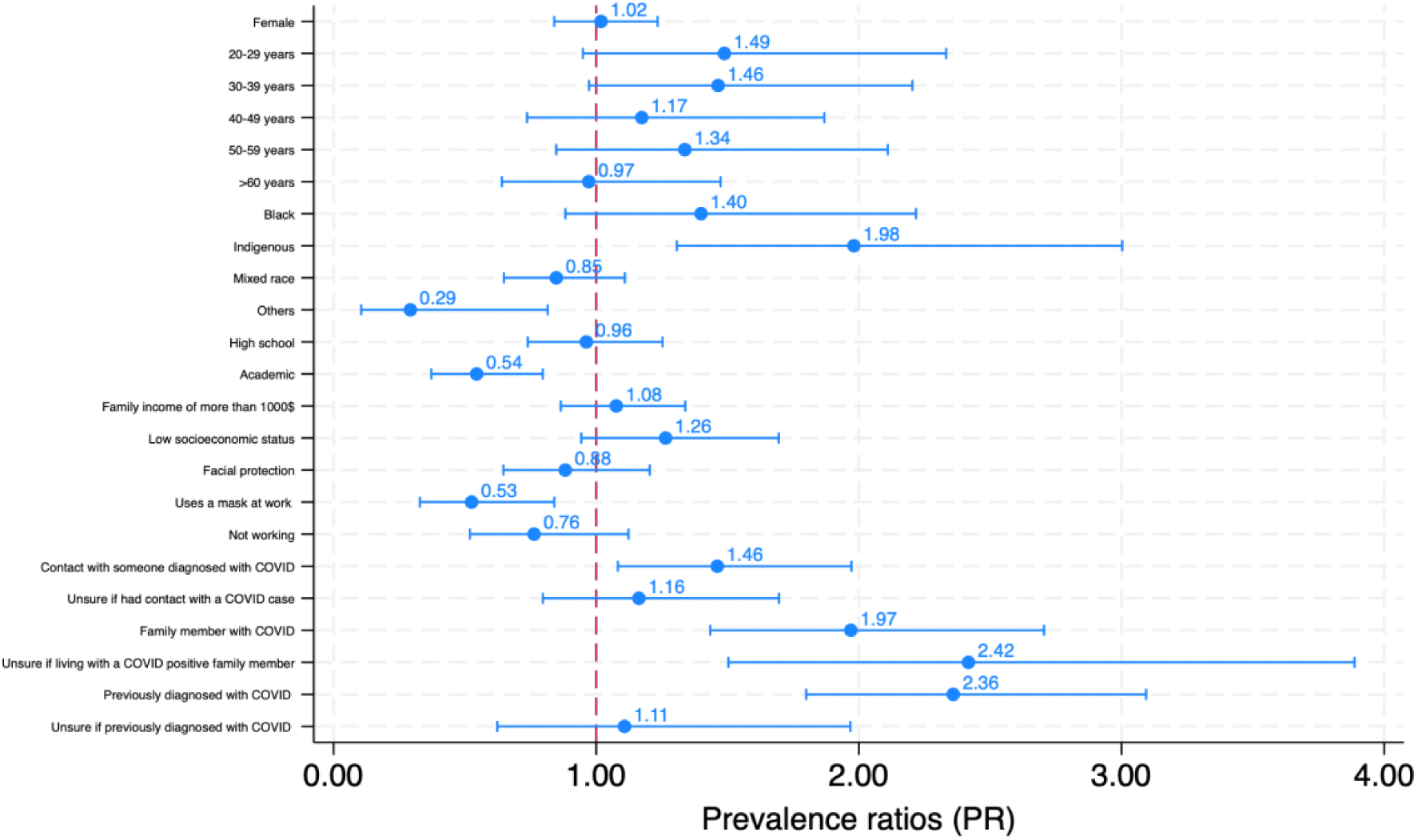
Prevalence ratios (PR) and 95% confidence intervals for factors associated with SARS-CoV-2 seroprevalence. The forest plot presents the results of a multivariable Poisson regression analysis evaluating associations between sociodemographic, occupational, and epidemiological factors and SARS-CoV-2 seropositivity. Variables with PR >1 indicate increased prevalence of seropositivity, while PR <1 suggests a protective effect.

## Discussion

After confirming the first SARS-CoV-2 cases, the Panamanian government swiftly imposed strict quarantine measures that lasted nearly six months^7^. The initial six-month quarantine in Panama imposed strict movement restrictions, limiting individuals to only two hours per day for essential activities, such as grocery shopping or visiting pharmacies. Women were allowed to go out three days a week, while men were permitted two^9^. On weekends, there was a complete lockdown, with no outdoor activities allowed, including walking pets^9^. Despite these efforts, the increase in seroprevalence observed in our study makes clear that these non-pharmaceutical interventions (NPIs) were not sufficient to completely halt virus transmission. At the worldwide level the pandemic continued to spread even after the introduction of vaccines and despite the use of NPIs, and Panama did not escape from this reality. In 2024 cases of SARS-CoV2-infections are still reported^12^.

We conducted two cross sectional SARS-CoV-2 serosurveys in the ten townships with the highest confirmed COVID-19 incidence in Panamá and Panamá Oeste. In the first round (n = 2,198), performed shortly after the initial epidemic wave, overall seroprevalence was 24.4 %. Seven months later, in a second round (n = 547), seroprevalence increased to 66.2 %. Although the decline in participant retention between rounds must be considered when comparing these estimates, the first round seroprevalence primarily reflects intense community transmission, whereas the second round estimate is likely driven by both ongoing infection and vaccine induced antibody responses demonstrating a sustained presence of antibodies over time.

The high seroprevalence detected during the first round of sampling indicates significant community spread, even in the context of prolonged lockdowns. One likely driver for this continued transmission is household exposure^18^. Indeed, living with a confirmed case of COVID-19 was a risk factor in our study. During the strict quarantine, most of the population was confined to their homes, with only a limited number of individuals allowed to leave for essential work or daily activities^19^. The high seroprevalence level observed highlights the challenge of controlling a highly infectious virus, even under stringent mobility restrictions.

The findings highlight the need for further epidemiological research to assess the impact of these NPIs on the intensity and progression of the epidemic in Panama. Future studies should focus on understanding how factors such as household size, employment in essential sectors, and adherence to quarantine measures influenced transmission patterns. Additionally, a more in-depth exploration of the roles that public transportation, overcrowded housing conditions, and economic necessities played, as observed in a study conducted in Lima, Peru, during 2021, could provide valuable insights.^20^.

One key driver of sustained SARS-CoV-2 transmission was household exposure, despite Ministry of Health protocols for managing infected family members. In our study, close contact with and living alongside a confirmed COVID-19 case increased risk. Conversely, households with no resident or family member diagnosed with SARS-CoV-2 had a significantly lower risk, indicating that the absence of intrahousehold infection acted as a protective factor in the subdistricts with the highest case incidence.

While the age groups 20–29 and 30–39 years did not show statistically significant associations with SARS-CoV-2 seroprevalence, the observed marginal associations could be partly explained by the sample size limitations. Nonetheless, these findings are consistent with studies from Brazil and Peru, where similar age groups were identified as having higher exposure risks to the SARS-CoV-2 seroprevalence^20,21^. In Panama, these age groups make up a significant proportion of the country’s economically active population^22^. 20 to 39 year olds are likely to be more exposed due to their reliance on public transportation and other forms of mobility.

The association between indigenous status and SARS-CoV-2 prevalence is likely multi-factorial. Indigenous populations often face socioeconomic challenges, including higher levels of poverty and limited access to healthcare, sanitation, and other essential services^23–25^. These barriers can complicate disease prevention and treatment, heightening vulnerability to infections like SARS-CoV-2. Additionally, social behaviors common in many indigenous communities, such as group gatherings, may increase the risk of virus transmission, especially in the absence of robust public health measures^24^. Populations with lower socioeconomic status often experience increased exposure to infectious diseases due to factors like overcrowded living conditions, reliance on public transportation, and limited access to healthcare and protective measures. These conditions may have contributed to the higher SARS-CoV-2 seropositivity observed in these groups.

On the other hand, higher educational accomplishment appears to serve as a protective factor against SARS-CoV-2 infection. Studies from Peru, Brazil, and Europe also found that individuals with more education tended to have lower disease prevalence, likely due to several factors^26–28^. Higher education enables greater health literacy, leading to a improved understanding and adherence to public health guidelines. It also enables access to more stable employment opportunities, such as telecommuting, which reduces the risk of exposure^29^. In Panama, many institutions as schools, universities and companies rapid transitioned to virtual modalities, helping to reduce contact and transmission during the pandemic. Our findings are consistent with studies from Switzerland, which indicate that the protective effect of telecommuting and education may vary depending on the context, such as the availability of reliable data and study design, which also found that the protective effect of telecommuting and education was significant to SARS-CoV-2 transmission^30^. In this context education played an important role in mitigating infection risk in Panama^31^.

The second round of the study, conducted seven months after the first, revealed that the proportion of seroconverted individuals far exceeded the proportion of seroreverted cases, demonstrating a sustained presence of antibodies over time. Importantly, antibody detection was secured using a test based on the N protein of SARS-CoV-2, which helped mitigate the potential confounding effect of vaccination, as the vaccines administered in Panama were primarily RNA-based platforms targeting the Spike protein. This approach allowed for a more accurate assessment of natural infection rates, minimizing the influence of vaccine-induced antibodies. These results provide valuable insight into the state of the Panama epidemic in late 2020 to mid 2021, suggesting a significant level of acquired immunity within the population, which is consistent with the country’s efforts to curb transmission and limit fatalities during the critical months of the pandemic.

The findings from this study should be interpreted with certain limitations in mind. First, the results may not fully reflect the broader Panamanian population, as the study was concentrated in ten townships with the highest incidence of SARS-CoV-2 in the provinces of Panama and Panama Oeste, potentially restricting the generalizability of the conclusions to other provinces and townships in Panama. Additionally, the substantial decrease in sample size during the second round of data collection impacted precision of later estimates may have introduced selection bias. Despite these limitations, the study employed a rigorous sampling design that enabled the generation of reliable estimates of SARS-CoV-2 transmission in the most affected areas during 2020. Additionally, the collection of data at two different time points allowed for the evaluation of seroconversion and seroreversion processes over a seven-month interval. These findings provide valuable evidence to inform public health strategies and contribute to the understanding of SARS-CoV-2 dynamics in Central America.

In conclusion, our study highlights the persistence of SARS-CoV-2 transmission in the regions studied in Panama during the second half of 2020, despite the implementation of strict lockdown measures. Household transmission, particularly within Indigenous communities emerged as a key driver of infection. Higher education was identified as a protective factor, possibly linked to greater access to information and better adherence to preventive practices. Although non-pharmaceutical interventions (NPIs), such as nationwide lockdowns, were implemented, our data do not allow for a direct evaluation of their effectiveness. Nevertheless, the findings provide a more detailed understanding of SARS-CoV-2 transmission dynamics in the provinces of Panama and Panama Oeste, specifically in the 10 townships selected for this study. These results offer valuable evidence to better understand the factors that may have influenced the progression of the pandemic in these regions and provide useful information for refining future public health strategies in similar settings.

## Data Availability

All data produced in the present study are available upon reasonable request to the authors

## Acknowledgement

We are deeply thankful we so many institutions and personas who make possible this research under such unique Public Health circumstances. We thank the social movement #TodoPanama for all the support provided during the first round of the study. We thank, Judy Meana, Governor of the Province of Panama and, Yazmin Colon de Cortizo first Lady of the Republic of Panama who make possible all coordination with local leaders and authorities. We thank local leaders and local authorities for all the support provided during the field investigation. We thank the National Police for providing care and help during the fieldwork investigation. We also thank all the volunteers who participated and worked in the field collecting data.

---

**Supplementary table 1.**
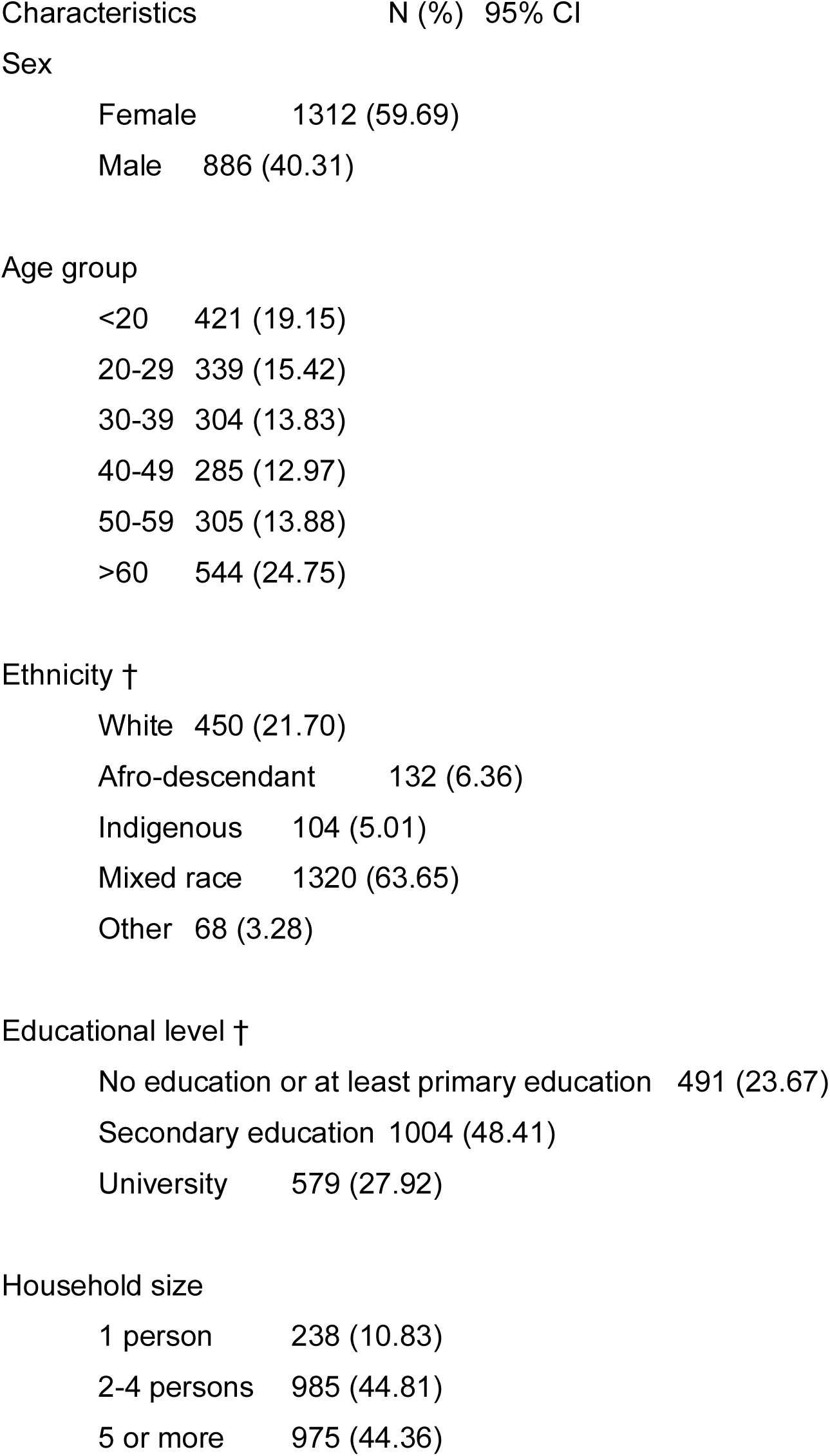
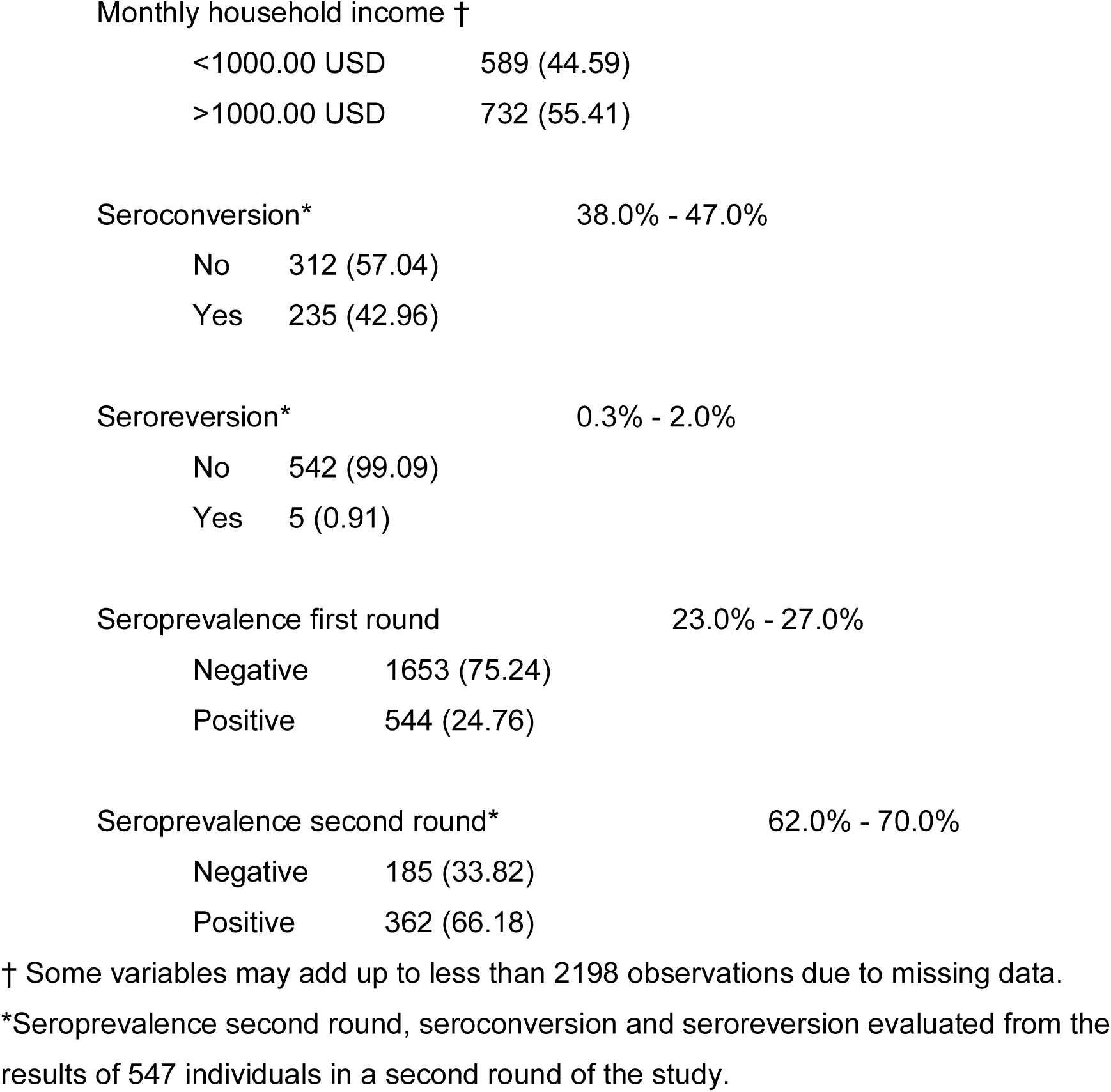
Sociodemographic characteristics and seroprevalence of SARS-CoV-2 in the Panamanian population, round 1 and 2 (n=2198)†.

